# Predicted COVID-19 fatality rates based on age, sex, comorbidities, and health system capacity

**DOI:** 10.1101/2020.06.05.20123489

**Authors:** Selene Ghisolfi, Ingvild Almås, Justin Sandefur, Tillmann von Carnap, Jesse Heitner, Tessa Bold

## Abstract

Early reports suggest the fatality rate from COVID-19 varies greatly across countries, but non-random testing and incomplete vital registration systems render it impossible to directly estimate the infection fatality rate (IFR) in many low- and middle-income countries. To fill this gap, we estimate the adjustments required to extrapolate estimates of the IFR from high- to lower-income regions. Accounting for differences in the distribution of age, sex, and relevant comorbidities yields substantial differences in the predicted IFR across 21 world regions, ranging from 0.11% in Western Sub-Saharan Africa to 0.95% for High Income Asia Pacific. However, these predictions must be treated as lower bounds, as they are grounded in fatality rates from countries with advanced health systems. In order to adjust for health system capacity, we incorporate regional differences in the relative odds of infection fatality from childhood influenza. This adjustment greatly diminishes, but does not entirely erase, the demography-based advantage predicted in the lowest income settings, with regional estimates of the predicted COVID-19 IFR ranging from 0.43% in Western Sub-Saharan Africa to 1.74% for Eastern Europe.

## 1. Introduction

Key policy decisions for COVID-19 containment hinge on the infection fatality rate (IFR) of this new disease. Data from the hardest-hit countries show that the IFR varies by sex, age and certain comorbidities, suggesting a method to extrapolate estimates to new contexts with limited data infrastructure [1].^1^ In this article, we combine recent estimates of the sex- and age-specific IFR from France with data on comorbidities conditional on death with COVID-19 in Italy to calculate the inverse: an IFR conditional on sex, age and comorbidity (cIFR). We apply these estimates to the distribution of sex, age, and relevant morbidities for 187 countries from the Global Burden of Disease (GBD) data set [8].^2^ Results reveal substantial differences across 21 world regions, with demographics-based IFR predictions ranging from 0.11% in Western Sub-Saharan Africa to 0.98% for High Income Asia Pacific. Despite the comparatively low IFR estimates our model predicts for the lowest income regions, these IFR estimates are appreciably higher than other recent estimates for the same areas [9].

We understand these predicted IFRs as lower bounds on mortality in low- and middle-income countries, since they are derived implicitly assuming access to French-level health care. We therefore calculate an adjustment for differences in health-system capacity in two steps. First, we extract COVID-19 mortality rates in French hospitals and condition them on age, sex, and comorbidities. Next, to account for the likelihood of higher fatality rates among hospitalized cases in under-resourced health systems, we incorporate regional differences in the relative odds of infection fatality from childhood influenza as a proxy for local capacity to treat viral respiratory illnesses. This adjustment greatly diminishes, but does not entirely erase, the demography-based advantage predicted in the lowest income settings, with regional estimates of the predicted COVID-19 IFR ranging from 0.43% in Western Sub-Saharan Africa to 1.74% for Eastern Europe. As a complement, we utilize hospitalization rates from France to estimate the probability of a severe COVID-19 morbidity, which is an object of independent interest.

## 2. Predicting the infection fatality rate conditional on age, sex, and comorbodities

In this section, we outline the calculation of our benchmark, predicted IFR conditional on age, sex and comorbidity status, starting from the IFR estimates by age and sex reported in [5] for France. The latter are, to our knowledge, the most recent peer-reviewed IFR estimates for COVID-19 which report variations for all age brackets and differentiate by sex. They are lower than earlier figures from [2], particularly among younger age groups, but are quite similar in the highest age brackets. As a first step, we define our target cIFR as the probability of dying (*d*) given infection from COVID-19 and the age, sex, and comorbidity status of patients. For the sake of notational simplicity, we denote probabilities conditional on infection (*I*), age(*a*), sex (*s*), and comorbidity status (*c*) using subscripts:

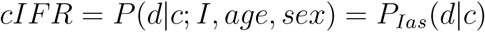

Applying Bayes’ rule we can recover the risk of dying conditional on age, sex and comorbidity by relating it to the ratio of comorbidity prevalence among COVID-19 fatalities relative to COVID-19 infections (conditional on age and sex) and age and sex-specific IFRs:

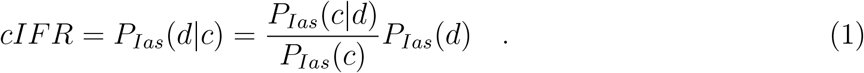

We now discuss how we measure each of these probabilities.

(1) *P_Ias_*(*c|d*) denotes the probability of comorbidity status given death of COVID-19, age and sex. We calculate this using the Italian Istituto Superiore della Sanità reports on the number of comorbidities conditional on COVID-19 death [10];^3^ relying on the assumption that this probability is independent of age and sex, *P_Ias_*(*c|d*) *≈ P* (*c|d, I*), which is supported by data from NYC.^4^ As shown in the appendix and justifying their combination, France and Italy are similar with respect to their estimated IFRs and comorbidity distribution.

(2) *P_Ias_*(*c*) denotes the presence of underlying conditions given infection, age and sex. We assume *P_Ias_*(*c*) ≈ *P* (*c|age, sex*) and take the reported probability of having any COVID-19-relevant comorbidity by age and sex in France from the GBD data set.^5^ Note that for simplicity we rely on an indicator for any COVID-19-relevant comorbidity, although the exact type, number, and combination of different diagnoses are likely to affect the cIFR.

(3) *P_Ias_*(*d*) denotes the sex and age-specific IFRs from [5], which come from France.

While these probabilities ((1), (2) and (3)) will vary across contexts as comorbidity rates vary, combining them via Bayes’ rule produces a conditional IFR (cIFR) that is purged – by virtue of conditioning – of variation in the underlying distribution of demographics and comorbidities. The core assumption underlying our calculations here is that our conditioning set is large enough to produce a relatively stable cIFR across countries.

We find the cIFR is an increasing and highly non-linear function of both age and comorbidity (Figure 1). For those without a comorbidity, the cIFR is effectively zero and flat up to the age of 50, and then increases roughly twenty-fold between 50–59 and 70–79 (from 0.01% to 0.17% for women and from 0.02% to 0.48% for men). With a comorbidity, the pattern is similar, but because the cIFR is already higher at younger ages, the age-gradient is less steep, roughly doubling the cIFR for each decade above age 50. The difference in the cIFR between patients with and without comorbidities is large but declines rapidly with age: a 30–39 year old is roughly 150 times more likely to die from COVID-19 if they have at least one comorbidity; at age 70 this ratio has decreased to roughly 10. Finally, the female cIFR is lower than the male cIFR for each age and comorbidity status.

**Figure 1:**
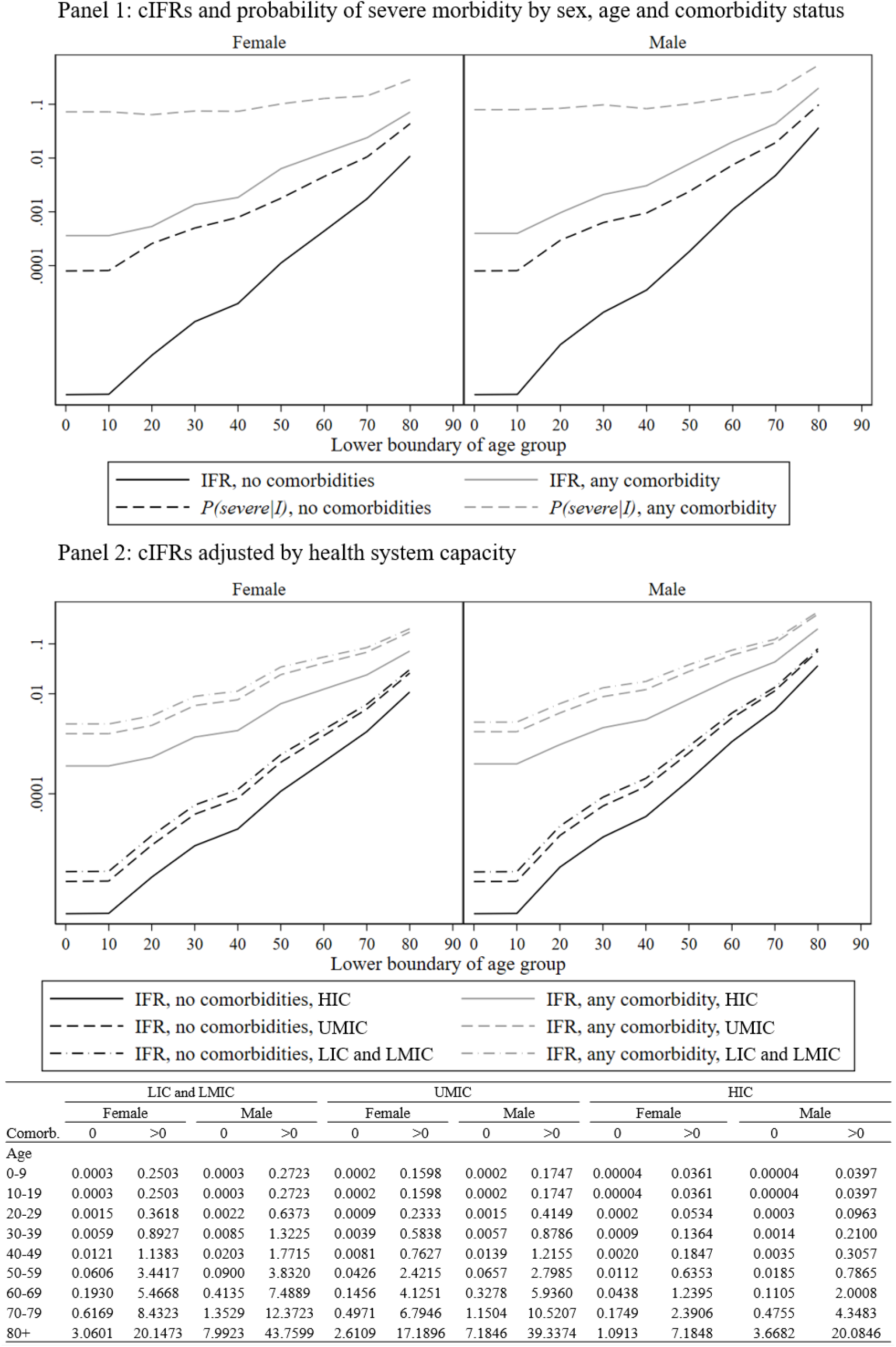
Panel 1 (graph): cIFR and probability of a severe case conditional on age, sex and comorbidity calculated from equations (1) and *(A.3)* (log scale). Panel 2 (graph and table): cIFR conditional on age, sex and comorbidity with and without adjusting for health system capacity by country income group, calculated from equations (1) and *(A.4)* (log scale in graph, levels in Table).

Figure 2 shows our main results, aggregated by 21 world regions. We find substantial variation in predicted IFRs across regions and countries – by a factor of 9 between the highest and the lowest. The variation is systematic, as low income regions have much lower predicted IFRs than high income regions. Demography is a key driver of these results: age distributions vary substantially across regions, with Sub-Saharan Africa and Oceania having the youngest and richer regions having the oldest populations. Regional variation in comorbidities also helps explain variation in predicted IFRs across regions: high-income regions display more comorbidities among the elderly than low-income settings, while the reverse is true among the young and middle-aged segments of the population. Finally, because the IFR is always lower for women than for men, variation in sex imbalances in the highest age brackets (biased towards women everywhere) also contributes to variation in the average IFR. A relatively low sex imbalance at ages 70 and above in some low income regions, such as South Asia, Carribbean, North Africa and the Middle East and Andean Latin America, increases the IFR in these places. On the other hand, female to male ratios among the elderly reach 3:1 in Eastern Europe, Sub-Saharan Africa and East Asia, decreasing expected mortality in these regions.

**Figure 2:**
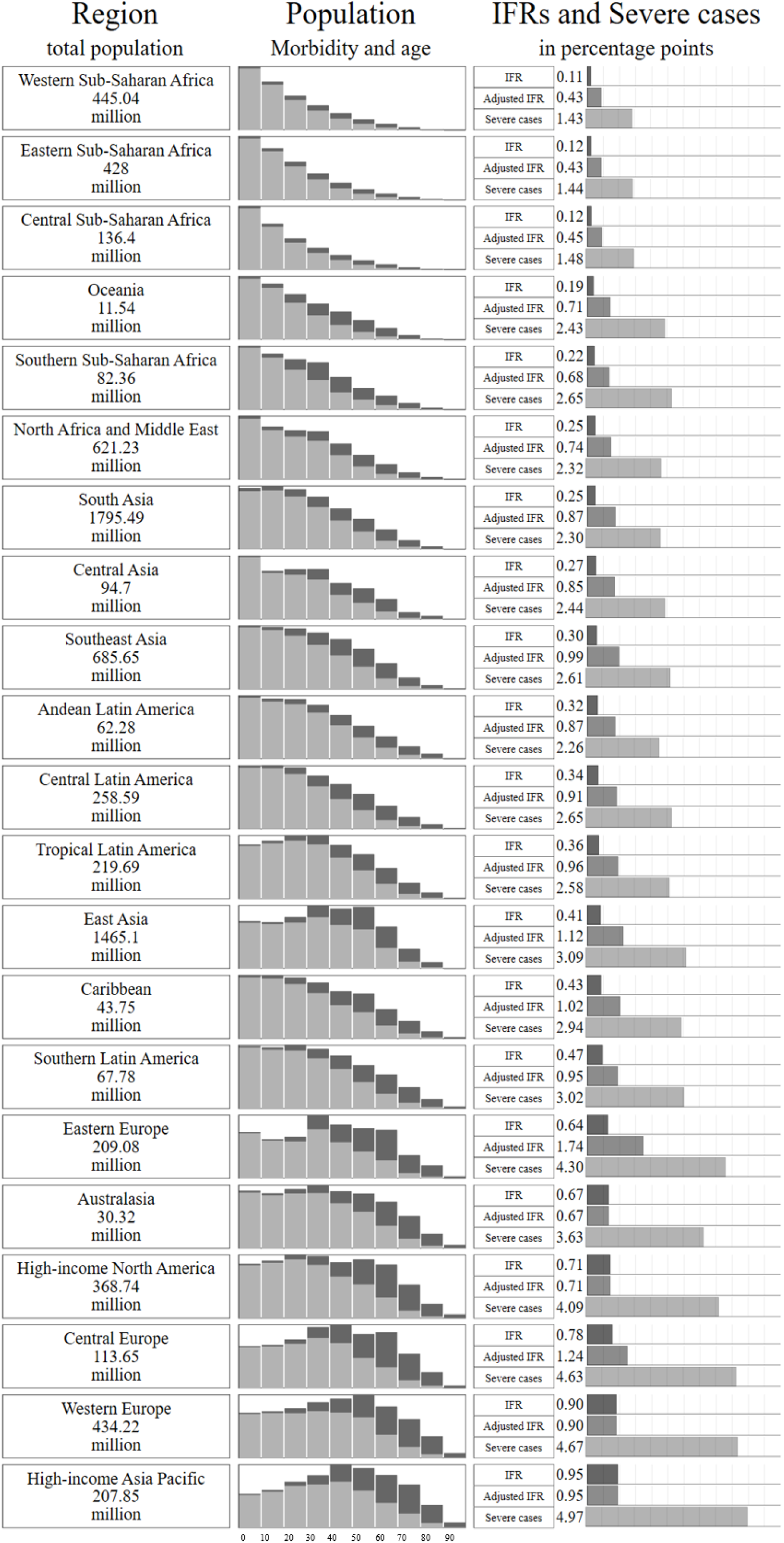
Infection fatality ratio by world region. Column 1 states total population in millions for each region. Column 2 reports population by 10-year age groups and by number of comorbidities (light grey – 0 comorbidities; dark grey – any comorbidity), the height of the graphs is proportional to the number of people in the most populous age group. Column 3 reports a) regional IFRs calculated as an average of the cIFRs by age, sex and comorbidity weighed by the proportion of the population in each age, sex and comorbidity group, b) regional IFRs adjusted for health system capacity (see section 4), and c) probability of a severe case (including deaths) given infection (see section 5)

## 3. Validating the predictions with serological studies from random samples

We can test the validity of our core assumption, i.e., that the cIFR is stable across contexts, by comparing our predicted IFRs to independently measured IFRs in a range of (mostly high-income) countries. For this exercise, we consider all studies reporting either IFRs or infection rates for populations with available COVID-19 fatalities, which were listed in the systematic review by [13] or retrieved through an online search on May 25. Out of the 30 studies selected in this way, seven studies measure infection rates by testing for seroprevalence of COVID-19 antibodies in population-based random samples. We judge this to be the best method of estimating infection rates, and thus IFRs, because random sampling is required to be truly representative, and antibody seroprevalence indicates all cumulative cases, whereas ‘swab’ tests only detect current cases. We thus compare our predicted IFRs first and foremost to the estimates in these seven studies. In a second step, we utilize all published IFR estimates in the comparison, including those which use convenience samples, adjusted Case Fatality Rates (CFRs), or ‘swab’ tests.

**Figure 3:**
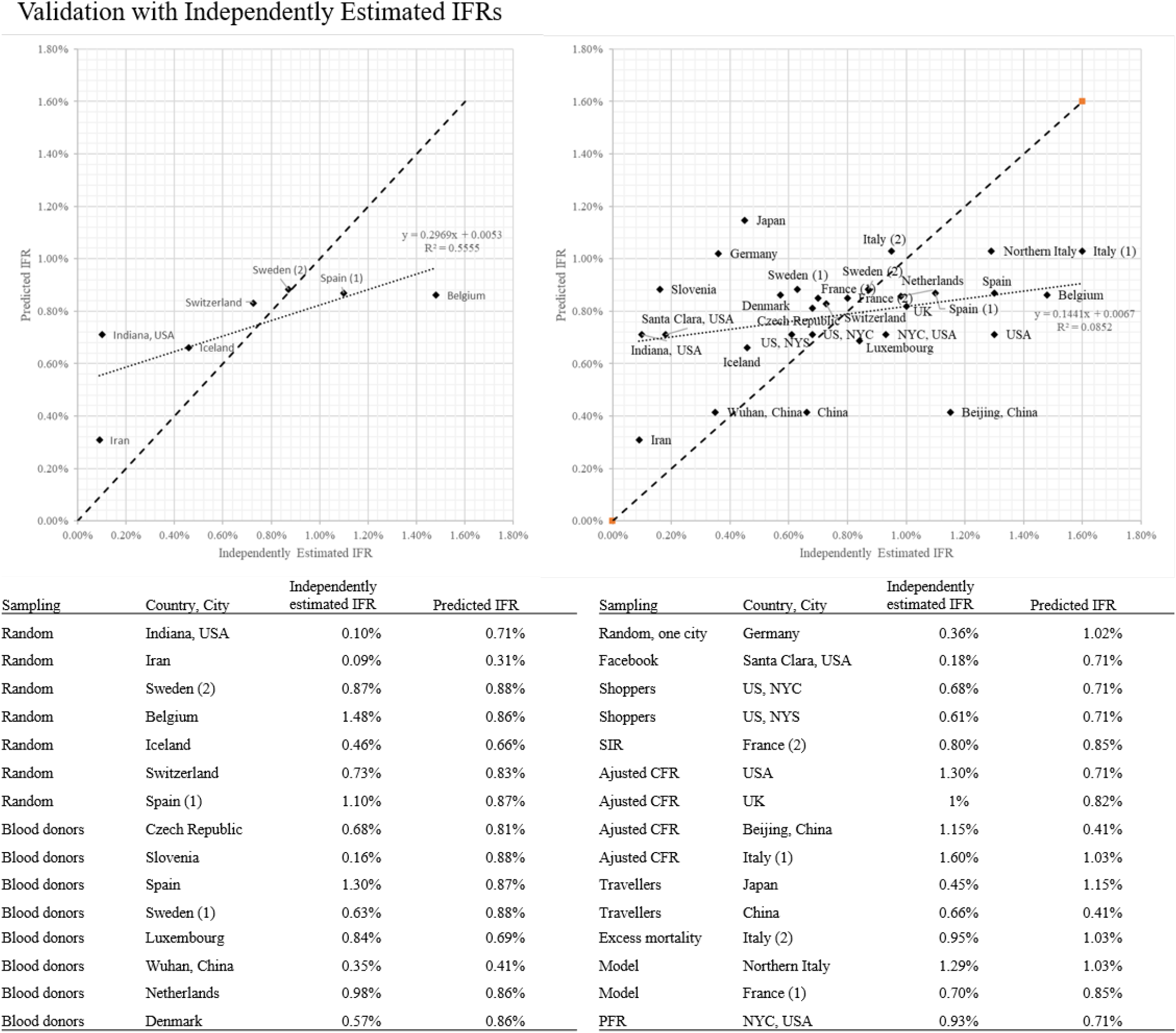
Validation with independently estimated IFRs. Left top panel: Random sample studies. Right top panel: All studies included in [13] or found through online search. Bottom panel: Table reporting studies included in the scatter plots.

The results are presented in the left top panel of Figure 3, where we plot the independent IFR estimates for the seven random-sampling studies in different countries on the horizontal axis against our predicted IFRs on the vertical axis. Comparing our estimates to these studies, we find an *R*^2^ of 0.56 and a slope of 0.30, indicating a good fit between our predictions and independent estimates. We note that, among these studies, Switzerland [14] and Sweden [15] are extremely close to the 45 degree line. The same holds for the estimates from Spain [16] and Iceland [17], which have been acknowledged to be well designed, randomized data collection efforts. Belgium [18], on the other hand, has a very high IFR relative to our predicted number, but it counts all suspect deaths in nursing homes as COVID-19 deaths^6^, yielding the highest IFR among the included studies. Other estimates further away from the 45 degree line are those from Indiana, USA [19] and from Iran [20]. In both cases, these are localized estimates, from one federal state in the USA and a single, heavily affected region in Iran. Thus, to properly compare the estimated IFRs to our predictions we would need to utilize local comorbidity, sex and age profiles rather than national ones.

Figure 3, right panel, reports the results from a comparison with all studies listed in [13], which include 9 studies which use modeling, CFR adjustments or excess mortality to predict the IFR, 12 studies with convenience samples drawn from blood donors, social media advertisements, recruitment in supermarkets or case studies in small cities, and 2 studies based on ‘swab’ tests from air travellers. The estimates displayed are much more noisy in this panel, including wide variations within single countries. Our predictions are meant to be nationally representative, and do not have as strong of a predictive power for IFR estimates that are not nationally representative, either from being highly localized in nature or non-random in their sampling. Further, our method assumes infection rates are uniform across demographics, which may not always hold at local levels. Nonetheless, our method does retain a positive correlation, albeit a low one, with even these measured IFRs. Representative sampling from non-OECD contexts will be necessary to confirm our core assumption of portable cIFRs holds in lower-income settings.

## 4. Adjusting for differences in health system capacity

We interpret our predicted IFR estimates as lower bounds on the true proability of dying from COVID-19 around the world, as data on fatalities come from countries with advanced health systems. Health system weaknesses in lower income settings are likely to diminish their demographic advantages. To account for this, we adjust our IFR estimates for health-system strength based on a region’s relative demonstrated capacity to prevent fatalities from viral pneumonias. We derive this adjustment from comparative regional IFRs (or, if unavailable, hospital case fatality rates) for seasonal-influenza-precipitated acute lower respiratory infection (ALRI) amongst children aged 12–59 months.

We chose this demographic to derive our health system capacity measure because restricting attention to this age bracket approximately purges the influenza IFRs of cross-country variation in the distribution of ages, comorbidities (as children under five have very low burdens of chronic diseases such as hypertension, kidney disease, or other conditions of organ degradation), and sex (as sex ratios under 5 years are more balanced than for older groups). With nearly equivalent age, sex, and comorbidity rates in this demographic, we take remaining cross-country variation in the IFR for seasonal influenza to be attributable principally to health-system capacity. We choose influenza ALRI as a proxy for COVID-19 as they are viral lower respiratory infections with overlapping symptoms. As with all viruses, neither is treatable with antibiotics, and influenza, despite heroic efforts, has no vaccine capable of conferring both broad-spectrum and long-term immunity. Influenza is also a pandemic disease with a well-researched global evidence base.

Normalizing the IFR for childhood seasonal influenza in high-income countries (HICs) to 1, we apply the ratio of these IFRs between regions to scale up our demography- and comorbidity-adjusted IFR predictions. Unfortunately, we lack country-level IFR estimates. However, [21] provide data from which childhood influenza IFRs can be inferred by World Bank income level for HICs and low and lower-middle income countries (LMICs). The ratio of the IFR for childhood influenza between LMICs and HICs from this data is 7.15. IFR estimates are not available for upper-middle income countries (UMICs), thus we rely on hospital case fatality rates, which are roughly double those of HICs. Taking these ratios as odds ratios rather than risk ratios (to maintain coherent probability bounds) we rescale the predicted cIFRs by region-specific adjustment factors to calculate a cIFR adjusted for regional health-system capacity (see Appendix A.1 for details).

Adjusting for health-system capacity increases the cIFR in poorer regions of the world by almost an order of magnitude (Panel 2 in Figure 1). At ages 60 and below, the cIFR is increased by a factor of 6–7 in LICs and LMICs and by a factor of 4–5 in UMICs. For older ages, the increase in the cIFR is less stark, but the adjusted cIFR is still 2–3 times as large as the unadjusted one. Adjusting for health system capacity thus both increases the cIFR at each age and co-morbidity status and flattens its age gradient. The reason for this is simply that the ratio of severe cases (as defined by hospitalization) to deaths is much smaller among older age groups in France, so the adjustment for heath system capacity has less impact for these groups. This pattern would be consistent with Intensive Care Units reducing COVID-19 mortality more for younger people than the elderly.

With this health-system adjusted cIFR in hand, we then recalculate the country-specific IFRs. The results are added to Figure 2. This health-system strength adjustment derived from childhood viral pneumonia treatment capacity starkly increases the predicted COVID-19 IFRs for the lowest income regions, nearly though not entirely erasing their demographic advantages. Eastern Europe is predicted to have particularly high IFRs, seeing as it is characterized by an aging population, high prevalence of co-morbidities at a given age, and low predicted health system capacity based on its income levels.

Our method of accounting for differences in health-system capacity is crude in that we currently only have indicative numbers for viral pneumonia by income group, rather than national-level adjustments. However, the wide gap in childhood viral pneumonia fatality rates of between 2- and 7-fold between income groups has implications for COVID-19 IFRs that are too large to ignore. Further research into how best to adjust IFR predictions to incorporate health system capacity is highly warranted.

## 5. Predicting severe infection conditional on age, sex and comorbidities

In addition to mortality, severe COVID-19 morbidity is also a concern as hospitals allocate scarce bed capacity and COVID-19 potentially crowds out care for other conditions. We use age and sex dependent hospitalization rates estimated by [5] for France as a proxy for severe morbidity and transform these into the probability of a severe case conditional on age, sex and comorbidity, again applying Bayes’ rule.^7^

These probabilities are larger by construction, as they include the probability of death as well as recovery from severe sickness through hospital care. Notably, the ratio of severe cases to deaths is higher for younger people, and thus the age-gradient is flatter for severe cases than deaths (Panel 1, Figure 1).

Aggregating these probabilities over the age, sex and comorbidity distribution in each country, we obtain the probability of a severe case for each country around the world. The probability of a severe case is 5 to 17 times higher than the predicted IFR and 2 to 5 times higher than the health system adjusted IFR (Figure 2).

While estimating the probability of a severe case is of independent interest, it can also be be utilized as a complementary approach to adjusting IFRs for local health system capacity. In poorer settings where sufficient beds are unavailable, or where capacity to prevent severe cases from ultimately deteriorating is limited, a large portion of severe cases may actually result in death due to suboptimal hospital care. The distance between our unadjusted IFRs and our severe case rate predictions will be crossed as health system capacity lowers from French/HIC levels to contexts which fully lack functioning hospitals.

## 6. Conclusion

Our results illustrate the possibility of predicting COVID-19 IFRs with a methodology that (a) uses information readily available for most of the world, (b) relies on parsimonious and transparent assumptions, and (c) appears broadly consistent with the limited set of IFRs generated from random COVID-19 testing. The adjustments by age, comorbidity, and health system capacity all appear important, especially in low income contexts: calculating IFRs without adjusting for comorbidity would yield estimates for Africa up to 30% lower than our benchmark predictions, and 60% lower than our predictions if adjusting for neither comorbidity nor health system capacity.

COVID-19 presents policymakers with difficult trade-offs. Many governments in low- and middle-income countries have resorted to severe lockdown measures which impose high costs. Recent estimates suggest COVID-19 mitigation efforts and the reallocation of healthcare resources in the developing world could contribute indirectly to over one million additional child deaths [22]. These unintended consequences must be weighed against the direct toll of an unmitigated pandemic. While our calculations including adjustments for health system strength still suggest slightly lower IFRs in the least developed economies than the in most advanced economies, our estimates are significantly higher than IFRs used in other recent COVID-19 forecasts for Africa [9], [2] and the middle-income world [2]. In the absence of widespread testing or reliable vital registration systems, transparent calculations of likely IFRs provide an important input into optimal policy design under extreme uncertainty, particularly as the pandemic expands into new geographies and/or a second wave of infections arrives.

## Data Availability

The analysis relies on the following, publicly available data sources.
Administrative reports from the Italian Istituto Superiore di Sanita:
https://www.epicentro.iss.it/coronavirus/sars-cov-2-decessi-italia
Administrative reports from NYC Health:
https://www1.nyc.gov/assets/doh/downloads/pdf/imm/covid-19-daily-data-summary-deaths-05162020-1.pdf
Global Burden of Disease data: http://ghdx.healthdata.org/gbd-2017
Supplementary material reported in Salje et al (Science, 2020):
https://science.sciencemag.org/content/sci/suppl/2020/05/12/science.abc3517.DC1/abc3517_Salje_SM.pdf

## Appendix A.

### Appendix A.1. Health system capacity adjustment

To obtain the cIFR adjusted by regional health system capacity, we first decompose the cIFR in countries in income group *j*, where *j* is either high-income countries (HICs), upper middle-income countries (UMICs) or lower middle-income countries (LMICs), into the probability of dying in or outside the hospital:

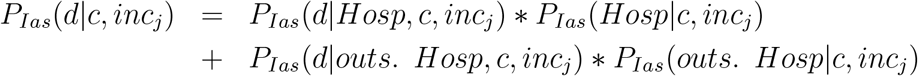

Above, *P_Ias_*(*Hosp|c, inc_j_*) is the probability (conditional on infection, age, sex and comorbidity status) of being hospitalized in a country in income group *inc_j_*, and *P_Ias_*(*d|Hosp, c, inc_j_*) is the probability of dying conditional on hospitalization in a country in income group *inc_j_*. *P_Ias_*(*outs. Hosp|c, inc_j_*) and *P_Ias_*(*d|outs. Hosp, c, inc_j_*) are defined in a similar manner. As above, we use the subscript *Ias* to indicate that COVID-19 infection, age and sex is always conditioned on. Throughout, we assume that the probability of a COVID-19 fatality occurring without first seeking hospital care is zero. Hence, we obtain

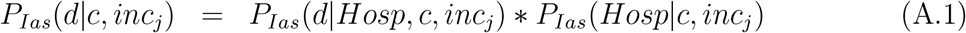

If the assumption that all deaths occur in hospital is violated, as is likely true in LMICs, our estimate of the cIFR adjusted by regional health system capacity is a lower bound.

We again apply Bayes’ rule to calculate the right-hand-side terms:

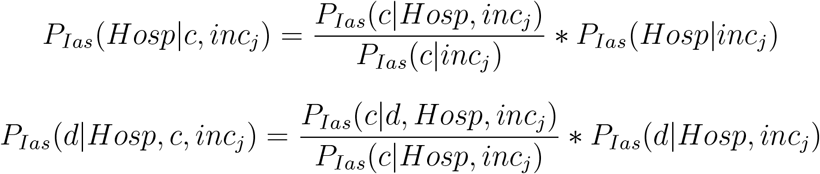

Substituting into (A.1), and simplifying we obtain

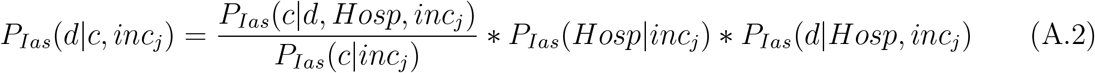

That is, the probability of death from COVID-19 conditional on age, sex, comorbidity and regional health system capacity equals the ratio of the prevalence of comorbidities among COVID-19 fatalities in hospitals to the prevalence of comorbidities among all COVID-19 infected multiplied by the probability of death from COVID-19 (in hospital) given the health system capacity in countries in income group *inc_j_*. We obtain these probabilities as follows:

1) *P_Ias_*(*c|d, Hosp, inc_j_*) and *P_Ias_*(*c|inc_j_*) are obtained in the same manner as in equation (1). That is, we use the Italian Istituto Superiore della Sanità reports on the number of comorbidities conditional on COVID-19 death. Even though we condition on hospitalization here, this is reasonable, since in any case, the data from Italy on prevalence of comorbidities is obtained from for the most part from hospital records. For *P_Ias_*(*c|inc_j_*), we make the same assumption as in the derivation of the cIFR, namely that the prevalence of comorbidities is independent of infection status.

2) To obtain *P_Ias_*(*Hosp|inc_j_*), we assume that the probability of hospital admission is the same across all income groups, *P_Ias_*(*Hosp|inc_j_*) = *P_Ias_*(*Hosp_j_*). The latter probability is again obtained from [23], which reports the probability for each age and sex to be admitted to a hospital in France with COVID-19 (p.36). This simplifying assumption is likely to be violated if, as we believe, holding constant the severity of the sickness, one is less likely to access a hospital in lower and upper middle income countries than in France. Given the direction of the violation, the resulting health-system adjusted cIFR is a lower bound for the true probability of death given infection in countries in income group *j*.

3) *P_Ias_*(*d|Hosp, inc_j_*): calculating the probability of dying from COVID-19 given hospitalization in countries in income group *j* is the main contribution of Section 4. It is calculated by multiplying the (age and sex-dependent) odds ratio of dying in hospital in France, which is obtained from [23] (p.37), by the health system capacity of countries in income group *j* relative to France. For LMICs, the latter is proxied by the ratio of under-5 infection fatality rates for childhood seasonal influenza between LMICs and France, 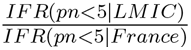, and for UMICs, it is proxied by the ratio of the under-5 hospital case fatality rate between UMICs and France, 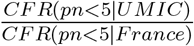. In either case, we denote the health system capacity of countries in income group *j* relative to France by *adj_incj_*. We thus write

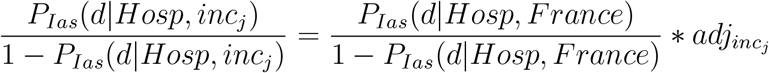

and solve for

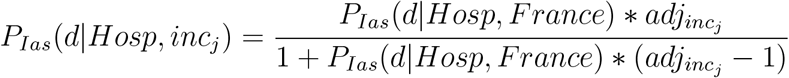

Substituting into (A.2) and simplifying, we obtain the health system adjusted cIFR:

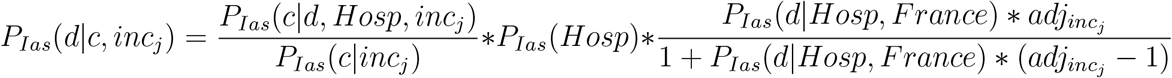

Under the assumption that all deaths happen in hospital, the final expression for the probability of dying from COVID-19 conditional on age, sex, comorbidity and health system capacity in countries in income group *j* simplifies to the product of the unadjusted cIFR times a scaling factor that varies by income group

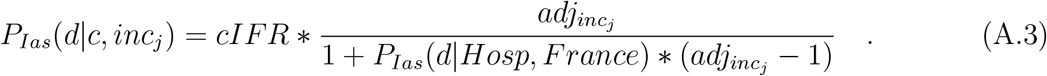

### Appendix A.2. Severe cases

We calculate the probability of a severe case (proxied by hospital admission in the French health system) conditional on age, sex and comorbidity status, i.e. *P_Ias_*(*severe|c*) ≈ *P_Ias_*(*hospital|c*). As with our benchmark cIFR calculations in the main text, we can calculate this from Bayes’ rule as follows:

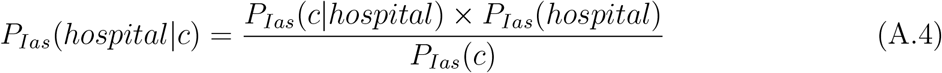

where *P* is the probability, *hospital* denotes hospital admission, *c* is the comorbidity status and *I* indicates infection. As above, we use the subscript *Ias* to indicate that COVID-19 infection, age and sex is always conditioned on. We review the two new terms in the numerator:

1) *P_Ias_*(*c|hospital*) denotes the comorbidity status given admission hospital. For the purposes of this calculation, we assume this probability to be equal to *P_Ias_*(*c|d*), the probability of having any comorbidity given death with COVID-19. This seems not implausible given data from the European CDC [24] that shows that at least 75% of ICU patients have a relevant comorbidity and data from China [25] that estimates a similar share among hospital admissions. The assumption implies that *P_Ias_*(*c|hospital, recover*) = *P_Ias_*(*c|d*), i.e. the comorbidity profile is the same between fatalities and the patients who were admitted to hospital and recovered.

We can speculate about the direction of the bias if this assumption is violated. If *P_Ias_*(*c ≥* 1*|hospital, recover*) *< P_Ias_*(*c ≥* 1*|d*), the prevalence of any comorbidities among fatalities would be higher than the prevalence of comorbidities among people who recovered after a hospital stay. The assumption would then lead us to overestimate *P_Ias_*(*severe|c*) for the population with comorbidities and underestimate it in the population without comorbidities. This would differentially impact our country-specific *P* (*severe|I*) according to the demography and comorbidity profile of the population. In countries with a healthier population per age and thus a higher number of healthy people overall our estimated probability would be a lower-bound for the true *P* (*severe|I*), while for countries with more comorbidities we would be overestimating the probability of a severe COVID-19 case. To confirm this is the most likely direction of the bias, we run a simulation assuming *P_Ias_*(*c ≥* 1*|hospital, recover*) = 75%^8^. Our simulation shows that indeed healthier countries at any given age (e.g. Peru, Japan) have a small increase of 3% in the probability of severe cases, while countries with more comorbidities (e.g. Lesotho, Swaziland, Botswana) show a decrease of 15% in their probability of critical cases – from 2.74% to 2.30%. This simulation not only confirms our speculation on the direction of the bias given our expected violation, but it also shows that the difference is relatively small in most countries.

2) *P_Ias_*(*hospital*) denotes the sex and age-specific probability of being admitted to hospital with COVID-19. This is reported by [23] (p.36).

Figure A.1, Panel 1 shows the IFRs estimated by [23] for France and by [12] for Italy. The IFR estimates in the two countries have very similar patterns, by age. We additionally report in the table the IFR by age profile and sex from [23], *P* (*d|I, age, sex*), that we use as an input into our calculation.

**Figure A.1:**
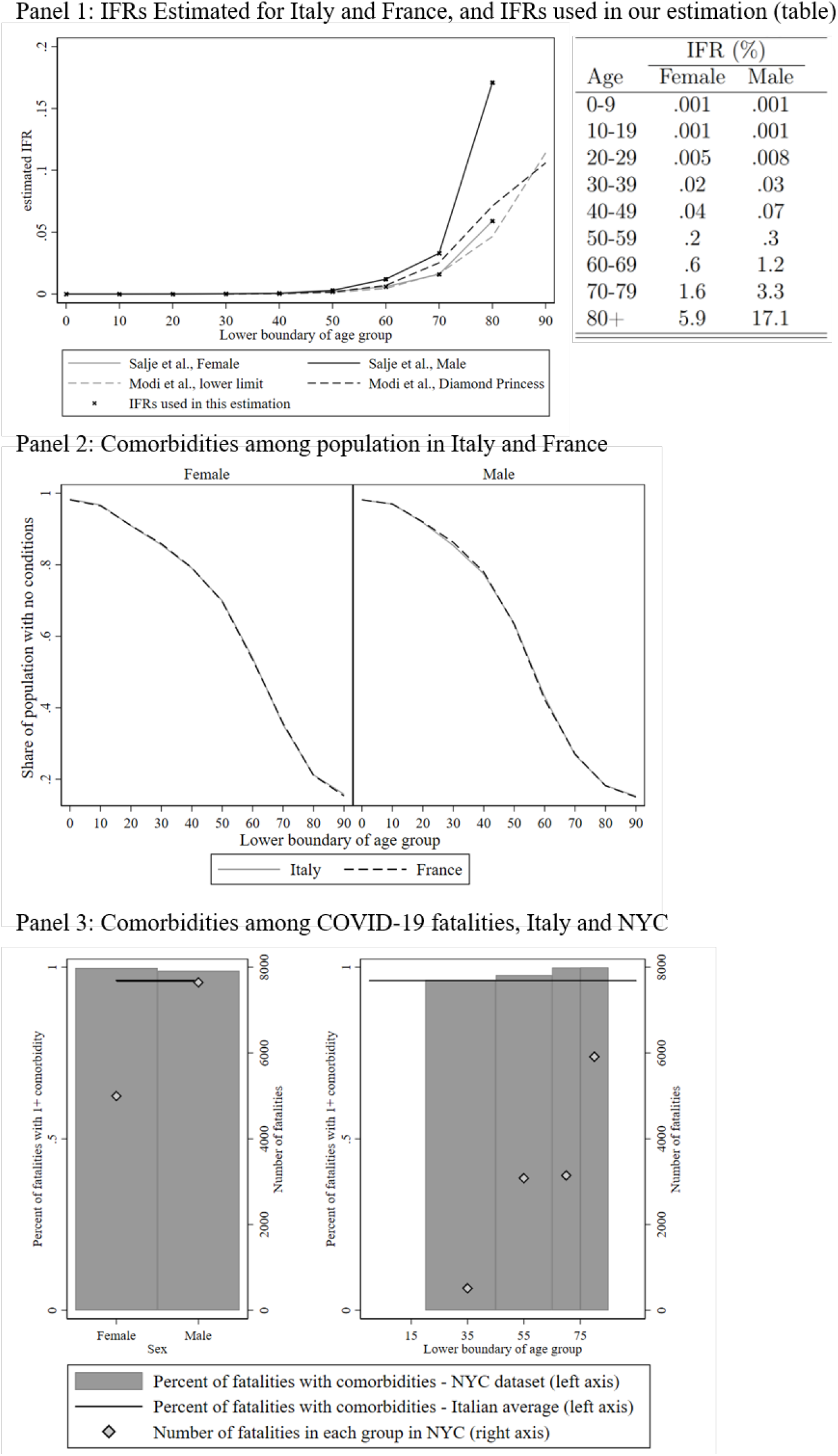
IFRs and morbidity profiles in the population of Italy and France, and comorbidity profiles of COVID-19 fatalities in NYC and Italy.

Figure A.1, Panel 2 shows the percentage of population with zero or any comorbidity by age and sex in France and Italy. The distributions are virtually identical. This gives us confidence in combining data from both countries for the estimation.

Figure A.1, Panel 3 shows the share of people who died with COVID-19 and any comorbidity in Italy (black line) and by sex and age group in New York City (bars and diamonds). The data is taken from Istituto Superiore della Sanità for Italy and New York City daily updates [7] on 16th May. Data for fatalities younger than 17 in NYC have been dropped in the age analysis as only nine deaths have been reported. The figure illustrates the very high presence of comorbidities among the COVID-19 fatalities for both sexes and in all age groups and the absence of a clear pattern by either of these variables.

Figure A.2 displays IFRs (in the first map) and IFR adjusting for health-system capacity by country income group (in the second map) for all the countries in the GBD data. Darker color indicates higher values, the scale is common in both maps, such that it is possible to compare the adjusted and the unadjusted IFR both within and across countries.

**Figure A.2:**
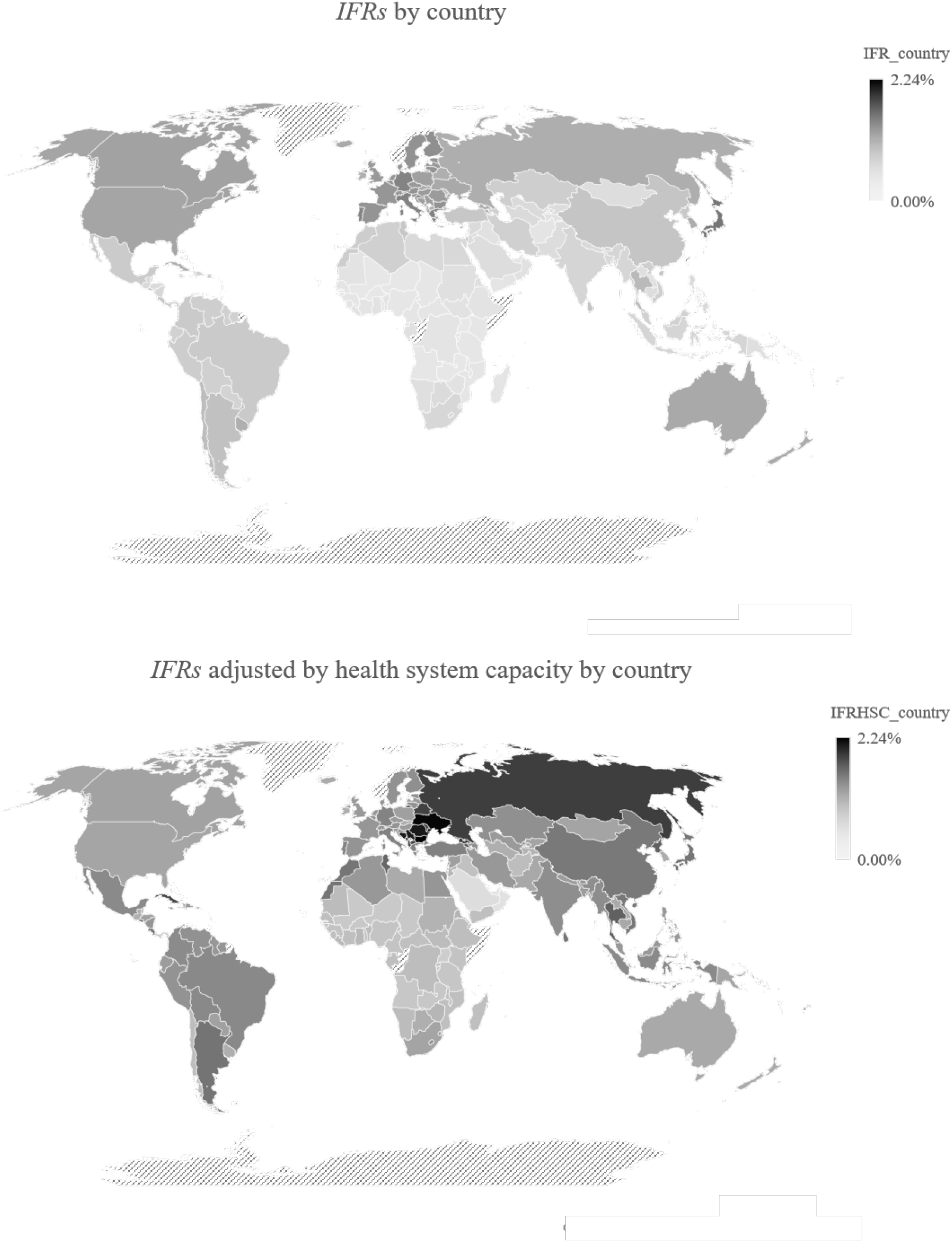
IFR with and without adjusting for health system capacity by country income groups. Darker color indicates higher values.

## Footnotes

1 [2] suggest large variation in mortality across countries. Early estimates indicated that age is an important factor for fatality of COVID-19 [3, 4]. Data from Italy and the UK also suggest an important role for certain comorbidities [5, 6]. This is further confirmed by data from New York City where a higher share of those who die with COVID-19 has a comorbidity, across age groups, than the general population [7].

2 The comorbidities considered relevant for COVID-19 by [8] are: cardiovascular diseases, chronic kidney diseases, chronic respiratory diseases, chronic liver disease, diabetes mellitus, cancers with direct immunosuppression, cancers with possible immunosuppression, HIV/AIDS, tuberculosis, chronic neurological disorders, sickle cell disorders

3 The most common diseases are hypertension (68% of deceased), type-2 diabetes (30.5%), ischemic heart disease (28.2% of deceased), atrial fibrillation (22.5%) and chronic renal failure (20.5%).

4 As shown in the Appendix, data from New York City indicate that among those who die from COVID-19, the share that has any comorbidity is stable across age groups and very similar for both sexes.

5 This assumption would be violated if the pool of infected systematically differs from the general population. Recent evidence from the US suggests that comorbidities are as present among the infected as in the general population [11]. Furthermore, data from Italy shows attack rates above 50% in some provinces. This, together with the absence of widespread immunity further supports this claim [12].

6 As reported in https://www.bbc.com/news/world-europe-52491210

7 See appendix A.2 for full derivation.

8 As reported in [24] [25]

